# Higher level of Quinolones residue in poultry meat and eggs; an alarming public health issue in Nepal

**DOI:** 10.1101/2022.09.03.22279573

**Authors:** Nabaraj Shrestha, Sundar Layalu, Serene Amatya, Samrat Shrestha, Shobha Basnet, Manash Shrestha, Upendra Thapa Shrestha

## Abstract

**Background:** With extensive use of antimicrobial agents in poultry production to prevent and treat diseases and as growth promoters, there has been an increase in antibiotic residue in the poultry meat and eggs. Quinolones are one of the highly prioritized, critically important drugs, whose residue in poultry can cause transmission of resistant zoonotic pathogens to humans. This study was conducted to assess the qualitative and quantitative residue of the quinolone in meat and eggs supplied to Kathmandu Valley, Nepal.

**Methods:** A standard questionnaire was used to identify the trend of antibiotic application in the poultry industry. Epicollect+ android application was used for the survey. A total of 120 meat samples and 120 egg samples were collected from the specified five sectors. Enzyme Linked Immuno Sorbant Assay was employed to screen antimicrobial residues in the samples. Only the samples with antimicrobial residues above maximum residual limit value were quantified with High Performance Liquid Chromatography.

**Result:** About 88.33% of chicken meat and 80% egg samples were positive for Quinolone residue. Three meat samples from Kathmandu had residues above the maximum residue level (MRL). One sample each from commercial and education sector showed Enrofloxacin residues while 1 sample from the health sector showed residues of both Enrofloxacin and Ciprofloxacin. The egg samples of Kathmandu, Bhaktapur and Lalitpur district contained 83.9%, 76.9% and 65% Quinolone residues respectively. While the eggs collected from household sector had the highest (100%) Quinolone residues than eggs from any other sectors followed by the lowest (66.6%) in eggs from the education sector (p=0.005).

**Conclusion:** A high level of Quinolone residue in the chicken meat and egg samples in the study indicates the imprudent and haphazard use of antibiotics which is a cause of emergence of antimicrobial resistant poultry pathogens alarming the potential transmission of resistance to human pathogens.

## BACKGROUND

Antibiotics have been widely used in the poultry industry as growth promoters, prophylaxis, and therapy, which are vital in producing meat and eggs (Phillips et al., 2004). The poultry industry and veterinarians have been using antibiotics to enhance growth and feed efficiency, reduce the bacterial load, and treat diseases (Donoghue, 2003; Darwish et al., 2013). However, the appropriate use of antibiotics has not been monitored by laws and guidelines in developing countries (Byarugaba, 2004). About 71% of veterinary drug sales are sold by self-prescription rather than by qualified registered veterinarians (Khatiwada & Acharya, 2013). Farmers use them without laboratory diagnosis, veterinary prescription, and supervision (Boisseau, 1993). The antibiotics such as aminoglycosides, tetracyclines, β-lactams, fluoroquinolones, macrolides, nitrofurans, sulphonamides, trimethoprim, etc., (Stolker & Brinkman, 2005; Lee et al., 2001) leave residues in foodstuffs of animal origin like meat, milk, and eggs (Darwish et al., 2013) and may exert different levels of toxicity on consumers (Fidel & Milagro, 2006). Above the maximum residual limit (MRL), the antibiotics residue can cause many disorders like immunopathological effects, carcinogenicity, mutagenicity, nephropathy, hepatotoxicity, reproductive disorders, bone marrow toxicity, and allergy (Nisha, 2008).

Recently antibiotics have been rampantly used as growth enhancers and treatments for diseases, resulting in multidrug resistance microbes such as *Escherichia coli, Salmonella spp, Campylobacter spp, and Staphylococcus aureus* of poultry origin. These resistant microbes are causing treatment failures and transmission of antibiotic-resistant zoonotic pathogens from animals to humans. The US-CDC thus recommends that antibiotics that are medically important to treating infections in humans should be used in food-producing animals only under veterinary oversight and only to manage and treat infectious diseases, not to promote growth (CDC, 2013). Furthermore, if use of antibiotics is indispensable, a withholding period must be observed until the residues are no longer detected or are below the MRL, depending upon the types of antibiotics used.

Fluoroquinolones are an important group of synthetic antimicrobials with broad-spectrum antibacterial activity, good absorption after oral administration, and extensive tissue distribution (Yang et al., 2005). Fluoroquinolones act via inhibition of DNA-gyrase, an essential enzyme in prokaryotes, abolishing its activity by interfering with the DNA rejoining reaction. (Yorke & Froc, 2000). They are widely used in the treatment and prevention of veterinary diseases in food-producing animals, and as growth-promoting agents. Enrofloxacin is the first specified Fluoroquinolones developed for a veterinary application, which belongs to the second generation of Quinolone antibiotics. Like other Fluoroquinolones, Enrofloxacin treats systemic infections, including urinary tract, respiratory, gastrointestinal, and skin infections (Tong et al., 2010). In addition, because of a very broad spectrum of activities against Gram-negative and Gram-positive bacteria and lower side effects, Enrofloxacin has also been widely used to treat some infectious diseases in pets, poultry, and livestock. However, Enrofloxacin residues may persist in the animal body, which may develop drug-resistant bacterial strains or allergies (Yan et al., 2011). Another alternative to Enrofloxacin is Ciprofloxacin. It is a significant metabolite of Enrofloxacin in animals and is also one of the most widely used Fluoroquinolones for treating urinary tract infections, respiratory tract infections, and chronic bacterial prostatitis (Ahmad et al., 2006). On the other hand, the Ciprofloxacin residues in livestock products may cause serious public health problems. Currently, awareness of residual antibiotics in animal-derived food is growing as their application increases in human and veterinary medicine. Therefore, more and more countries have set the MRLs and withdrawal periods for fluoroquinolones (Huet et al., 2006; Lu et al., 2006). For example, an MRL of 100μg/kg has been recommended for the sum of Enrofloxacin and its major metabolite, Ciprofloxacin, in animal muscles (Huet et al., 2006).

In contest of Nepal, the antibiotic application to poultry has increased along with the increasing consumption of eggs and poultry meat. However, limited research has been conducted regarding assessment of drug residue in poultry products for food safety in Nepal. Therefore, this research was carried out to quantify the residue level of the most widely used antibiotic in poultry meat and eggs consumed by people in Kathmandu valley. The outcomes of this research will provide supportive empirical evidence to inform a national policy on the prudent use of antibiotics in livestock and poultry. In addition, poultry practicing veterinarians, feed industry consultants, producers, consumers, and other stakeholders will receive a positive understanding of the imprudent use of antibiotics. Lastly, it is hoped that this research will also address the issues of One Health.

## METHODS

### Study site

This study was conducted in the three districts of Kathmandu valley (i.e. Kathmandu, Bhaktapur, and Lalitpur). A questionnaire was developed and pre-tested before using it in the field to identify the trend of antibiotic application in the poultry industry. Epicollect+ android application was used for the survey.

### Patient and public involvement

Nepalese poultry practicing veterinarians/consultants, non-vet feed consultants, drug distributors, layer farmers, broiler farmers were selected as the respondents to the questionnaire. Five sectors were identified within the three selected districts based on place for policy development, consumer, and potential zone at risk (Figure 1). The results of the study were disseminated among those respondents at seminars and group discussions organized at local settings.

**Figure 1.**
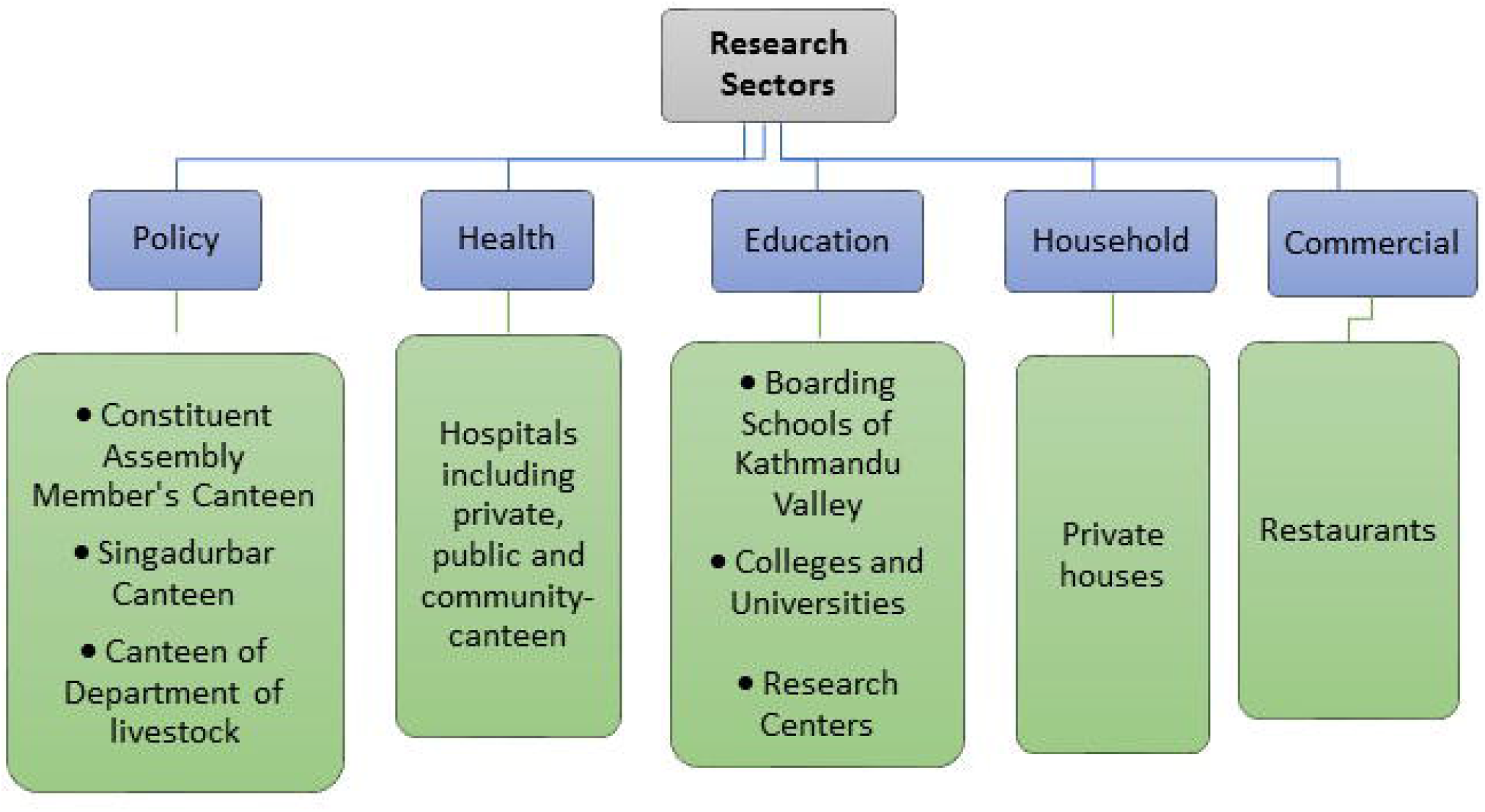

### Consent and data collection

Written informed consent was obtained from each of the participants involved in the research study using a standard consent form.

### Sample type and collection

Meat and egg samples were collected from the specified five sectors representing: the commercial, household, policy, education, and health sector. A total of 120 meat samples and 120 egg samples were collected (Table 1). From each of the sectors, 100 grams of meat sample and one egg sample were taken. Attention was given to discourage duplication such that no meat and egg sample belonged to the same farm and same batch.

**Table 1:**
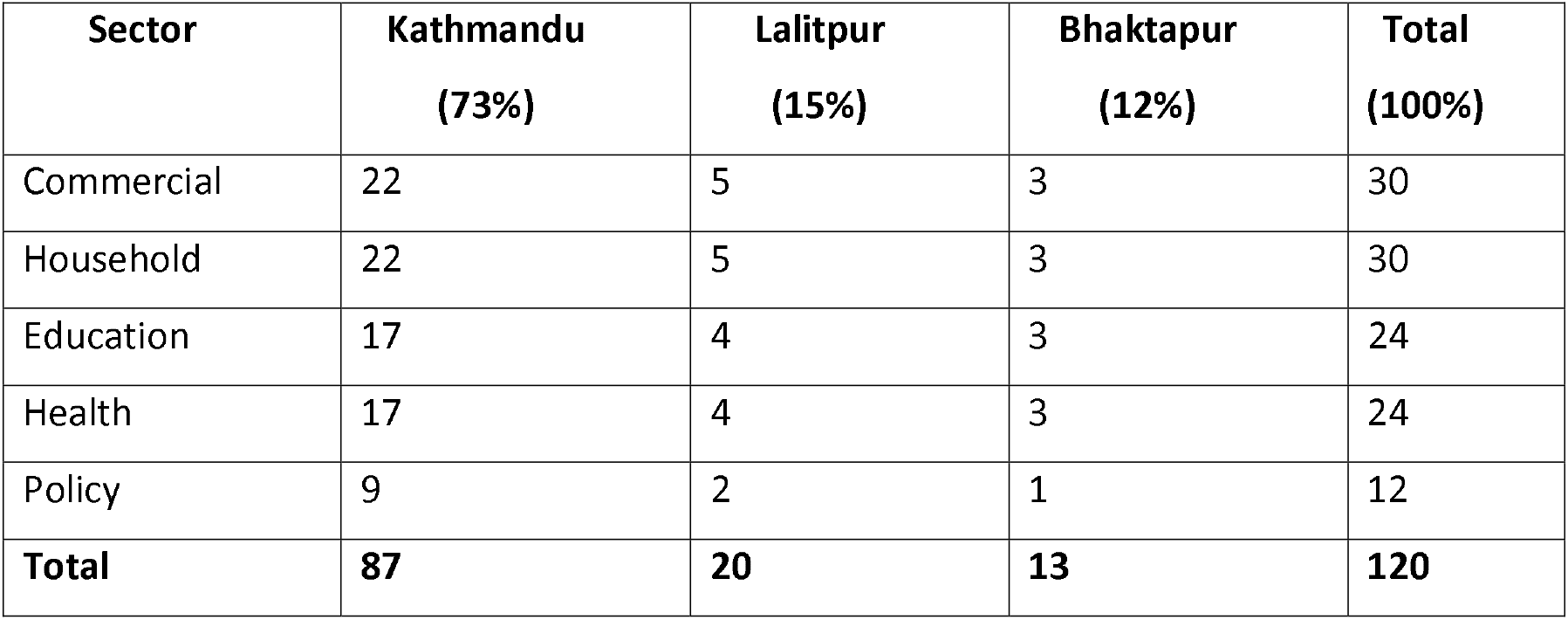
Tabular sketch of research sectors.

### Analysis of samples

Enzyme Linked Immuno Sorbant Assay (ELISA) was employed to screen antimicrobial residues in the meat and egg samples. Only the samples with antimicrobial residues above MRL value were quantified with High Performance Liquid Chromatography (HPLC). All laboratory procedures were performed at the Zest Laboratory, Balkot, Bhaktapur, Nepal.

### ELISA Test

Quinolone ELISA test kits were brought from the Shenzhen Lvshiyuan Biotechnology Co., Ltd, China and was performed for the qualitative analysis following the manufacturer’s guidelines. The concentration range (ng/ml) obtained from comparing the sample’s average Optical Density (OD) value with that of the standard solution was used to determine the presence or absence of Quinolones.

### High Performance Liquid Chromatography (HPLC)

The HPLC was performed to assess the Quinolones residues according to the methods mentioned in The United State Pharmacopeia, The National Formulary 2015, Volume 2 A (USP, 2015). A standard was prepared with 0.05% w/v solution of Enrofloxacin and Ciprofloxacin, each in water. The samples were prepared following the method of sample extraction for the ELISA screening test. A mobile phase was prepared with a mixture of 87 volumes of 0.025 M Phosphoric acid, previously adjusted with Triethylamine to pH 3.0±0.1 and 13 volumes of Acetonitrile, and filtered through 0.45μm membrane filter. A stainless-steel column 25 cm ×4 mm, packed with octadecylsilane (C18) bonded to porous silica (5 μm) was used with temperature of 30°±1°C. The injection volume was 20 L and flow rate was maintained at 1.5 ml/min. The residue was detected at 278 nm.

### Statistical Analysis

Data entry was managed in Microsoft Excel 2016, and the statistical analysis was done using R Ver. 3.4.0 (R core team, 2017). Poultry meat and eggs correlation among three districts and five sectors were carried out using Fisher’s exact test to identify antibiotics residue in specific district and sector.

## RESULTS

### Result of the questionnaire survey

In total, 105 respondents were surveyed through the questionnaire. Major antimicrobials identified in the poultry industry were Colistin sulphate, Enrofloxacin, Levofloxacin, Ciprofloxacin, Sulphonamides, Trimethoprim, Neomycin, Doxycycline, Tylosin, Amoxycillin etc. Similarly, minor antimicrobials identified were Tetracycline, Amikacin, Chlortetracycline, Oxytetracycline, Tiamulin, Cephalexin, Norfloxacin, etc. Among these antimicrobials, Colistin, Enrofloxacin, and Ciprofloxacin were the major antimicrobials used for treating broilers. Likewise, Doxycycline and Neomycin in combination was the most widely used antimicrobial in broilers for disease prevention, and BMD (Bacitracin Methylene Disalicylate) and Chlortetracycline were used as feed supplements. Similarly, Colistin sulphate, Ciprofloxacin and Tylosin were the major antimicrobials used in layers as a treatment. Likewise, Doxycycline, Neomycin and Tylosin were the major antimicrobials used in layers as prevention, and BMD, Chlortetracycline and Colistin were the three major antimicrobials used in layers as growth promoters. In addition, Colistin sulphate, Enrofloxacin and Doxycycline were the three major antimicrobials that the drug distributors sold in the highest amount.

### District-wise analysis of meat samples

In a total of 120 meat samples, 87 were from Kathmandu of which 77 samples (88.5%) were positive, 20 were from Lalitpur of which 17 samples (85%) were positive, and 13 were from Bhaktapur of which 12 samples (92.3%) were found positive for Quinolones antibiotics (Figure 2). There was no significant variation (p>0.05) in occurrence of antibiotic residue on meat samples across the three districts of Kathmandu valley.

**Figure 2.**
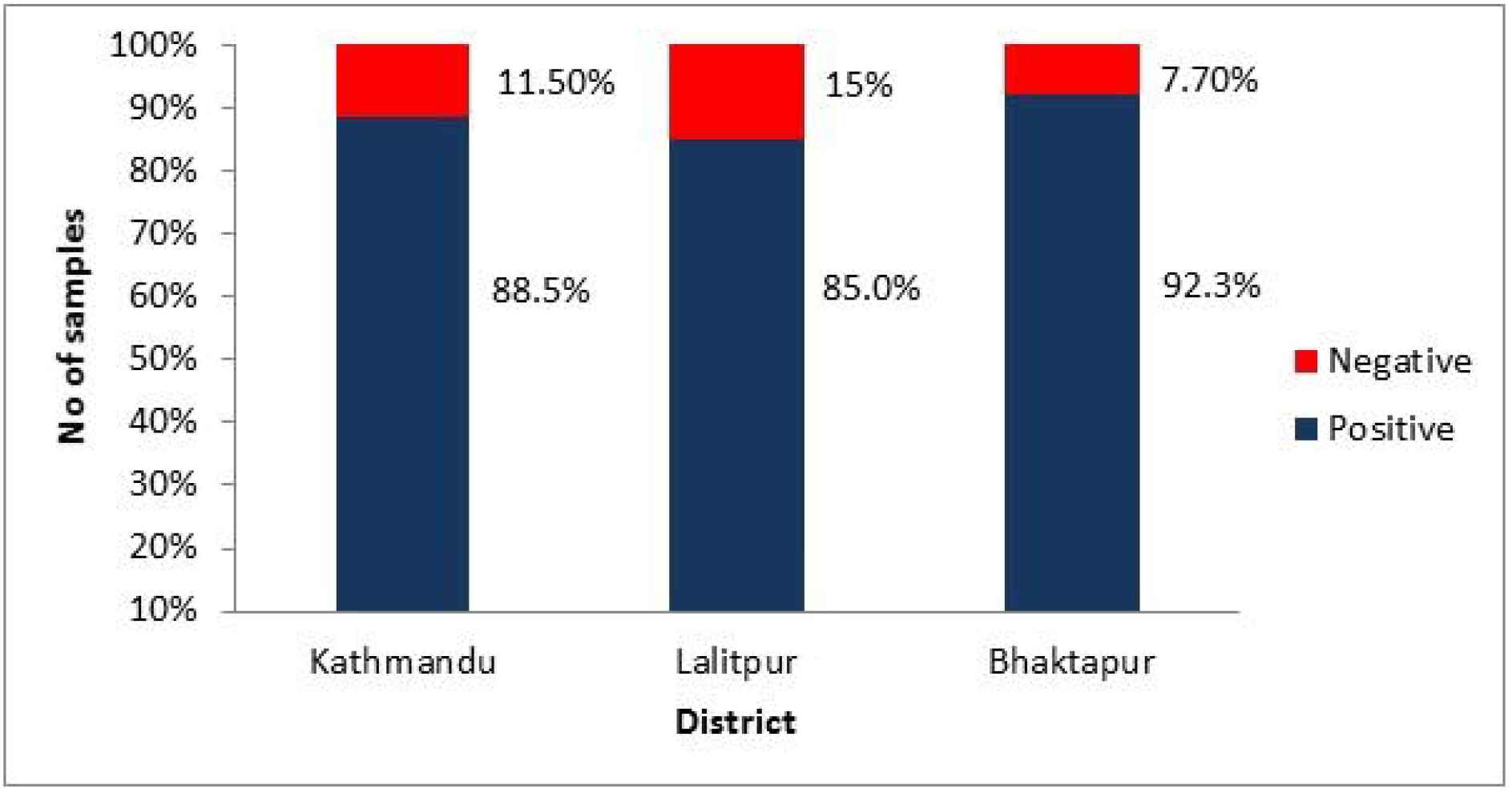

### Sector-wise analysis of meat samples

Out of 120 meat samples; 30 were from commercial sector of which 24 samples (80%) were positive, 30 were from household sector of which 28 samples (93.3%) were positive, 12 samples were from policy sector of which 9 samples (75%) were positive, 24 samples were from education sector of which 21 samples (87.5%) were positive, and 24 samples were from health sector of which all 24 samples (100%) were found to be positive for Quinolones antibiotic (Table1, Figure 3). There was no significant difference (p>0.05) in occurrence of antibiotic residue in meat samples among the various sectors.

**Figure 3.**
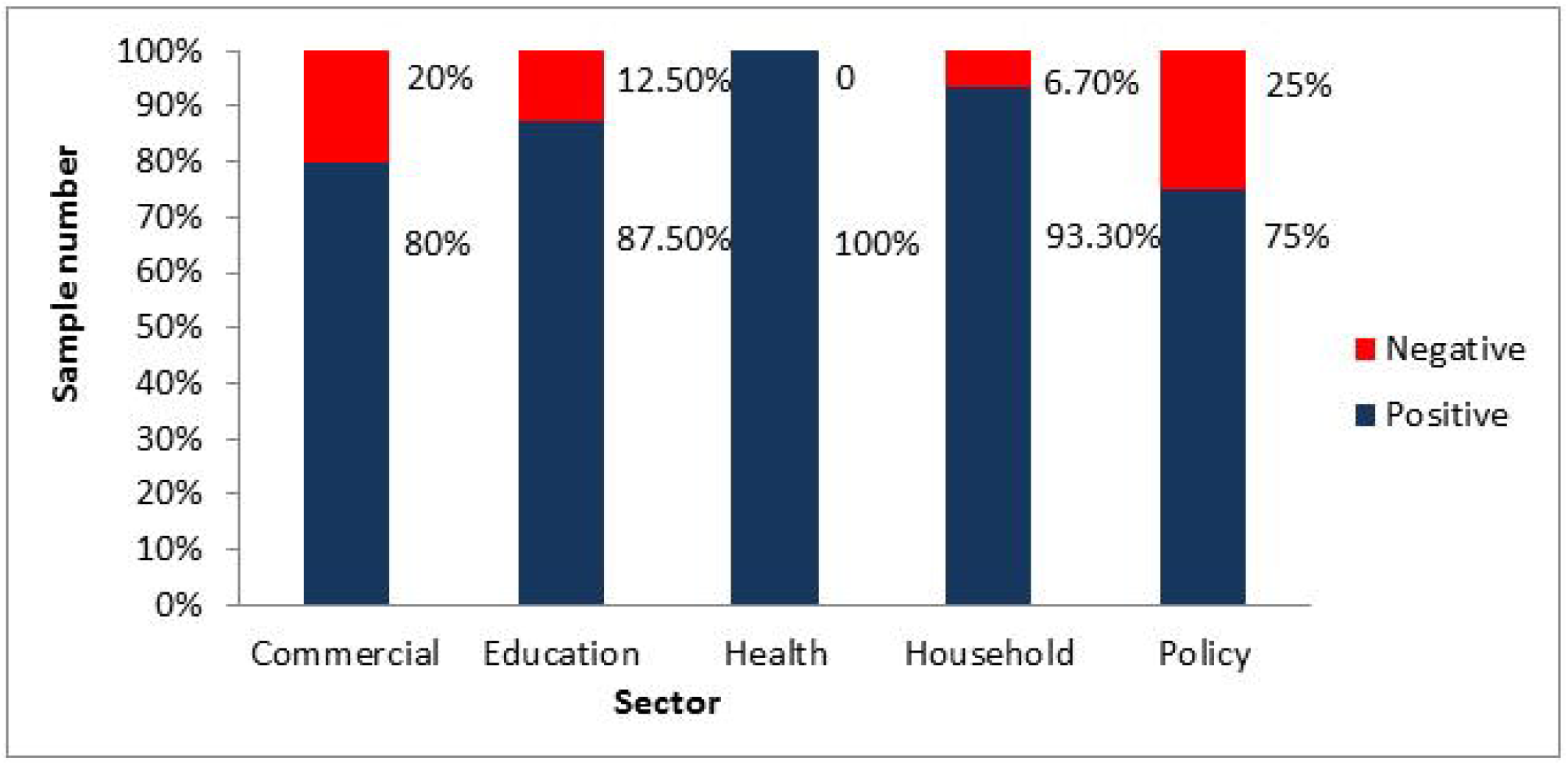

### Qualitative analysis of Quinolones residue in meat

Out of 120 meat samples, 106 (88.33%) samples were found to be positive for the residues of Quinolones. Out of positive samples, 9 samples (8.49%) were found to have residues above the MRL (i.e. 100μg/kg) (Table 2).

**Table 2:**
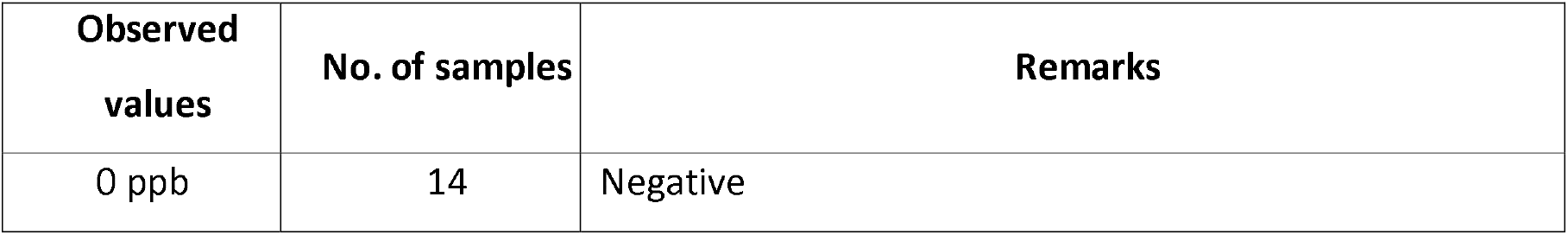
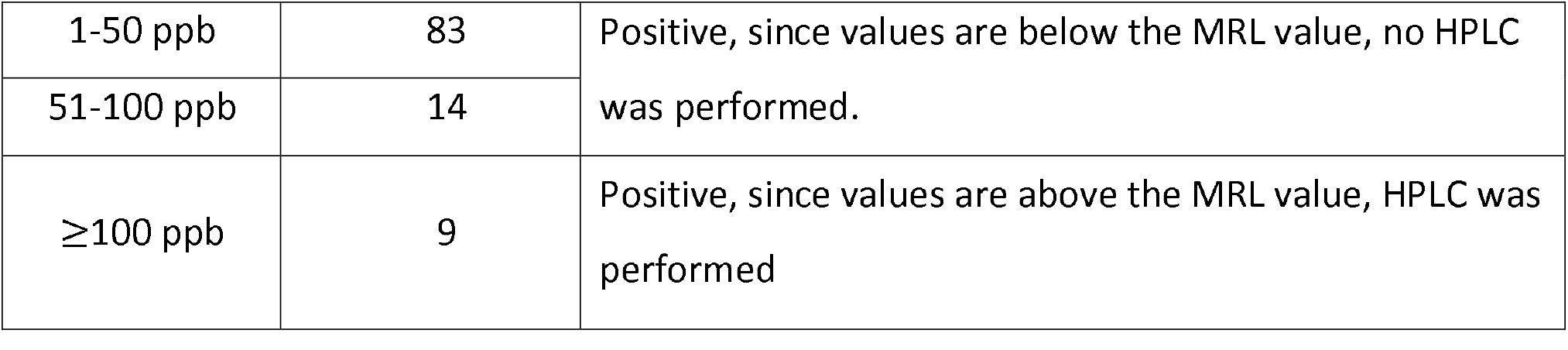
Observed values of the positive meat samples.

| **Observed**<br>**values** | **No. of samples** | **Remarks** |
| --- | --- | --- |
| 0 ppb | 14 | Negative |
| 1-50 ppb | 83 | Positive, since values are below the MRL value, no HPLC was performed. |
| 51-100 ppb | 14 |  |
| ≥100 ppb | 9 | Positive, since values are above the MRL value, HPLC was performed |

### District-wise analysis of antibiotic residue in egg samples

Out of the total 120 egg samples; 87 were from Kathmandu of which 73 samples (83.9%) were positive, 20 were from Lalitpur of which 13 samples (76.9%) were positive, and 13 were from Bhaktapur of which 10 samples (65%) were found positive for Quinolones antibiotics (Figure 4). There was no significant variation (p>0.05) in occurrence of antibiotic residue on egg samples among the three districts of Kathmandu valley.

**Figure 4.**
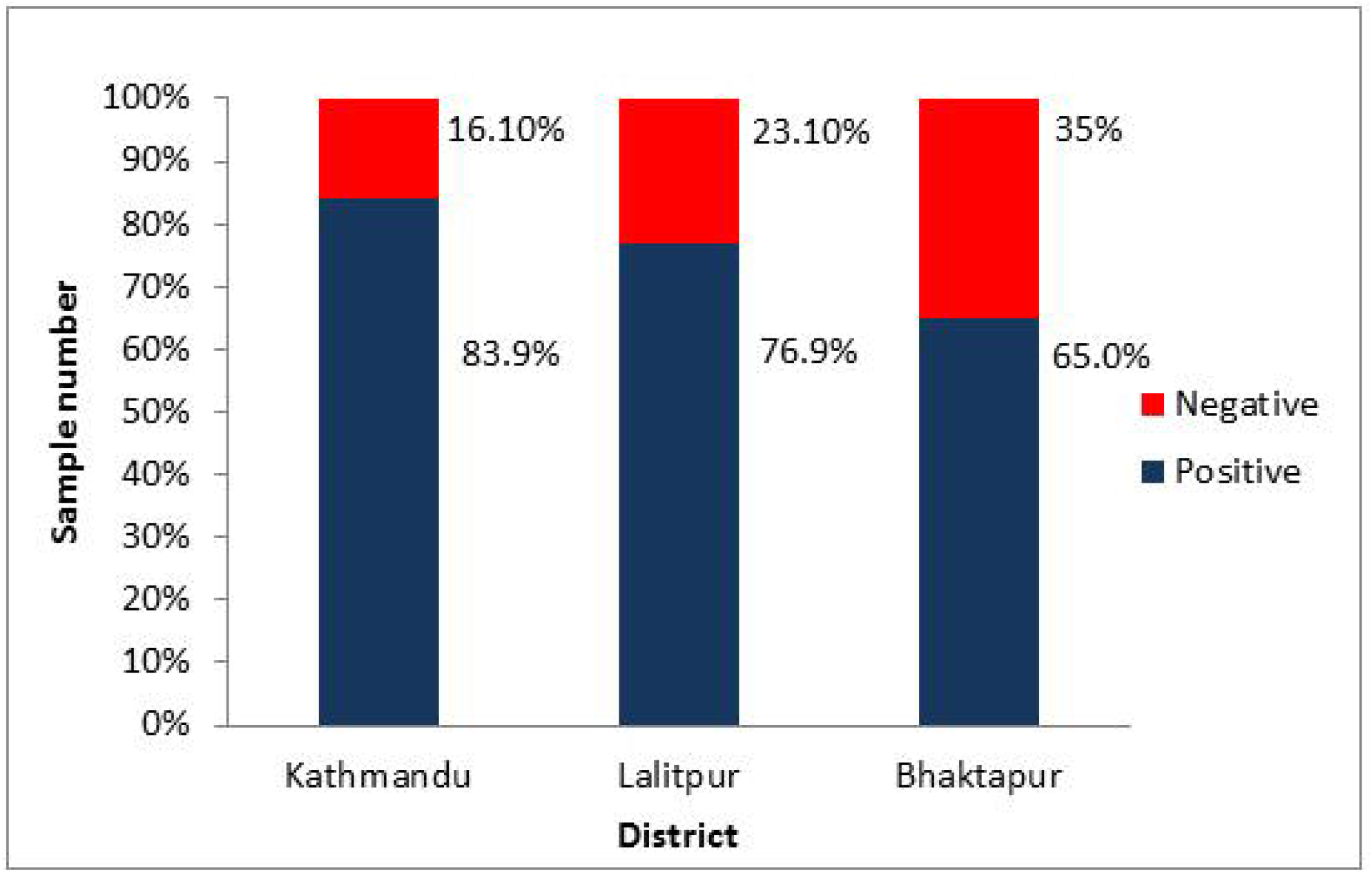

### Sector-wise analysis of antibiotic residue in egg

Out of 120 egg samples; 30 were from the commercial sector of which 22 samples (73.3%) were positive, 30 were from the household sector of which all 30 samples (100%) were positive, 12 samples were from the policy sector of which 10 samples (83.3%) were positive, 24 samples were from the education sector of which 16 samples (66.6%) were positive, and 24 samples were from the health sector of which 18 samples (75%) were found to be positive for Quinolones antibiotic (Figure 5). There was significant variation (p<0.05) in occurrence of antibiotic residue in egg between household and that of commercial, education and health sector. There was no significant difference (p>0.05) in antibiotic residue between policy sector and that of any other sector.

**Figure 5.**
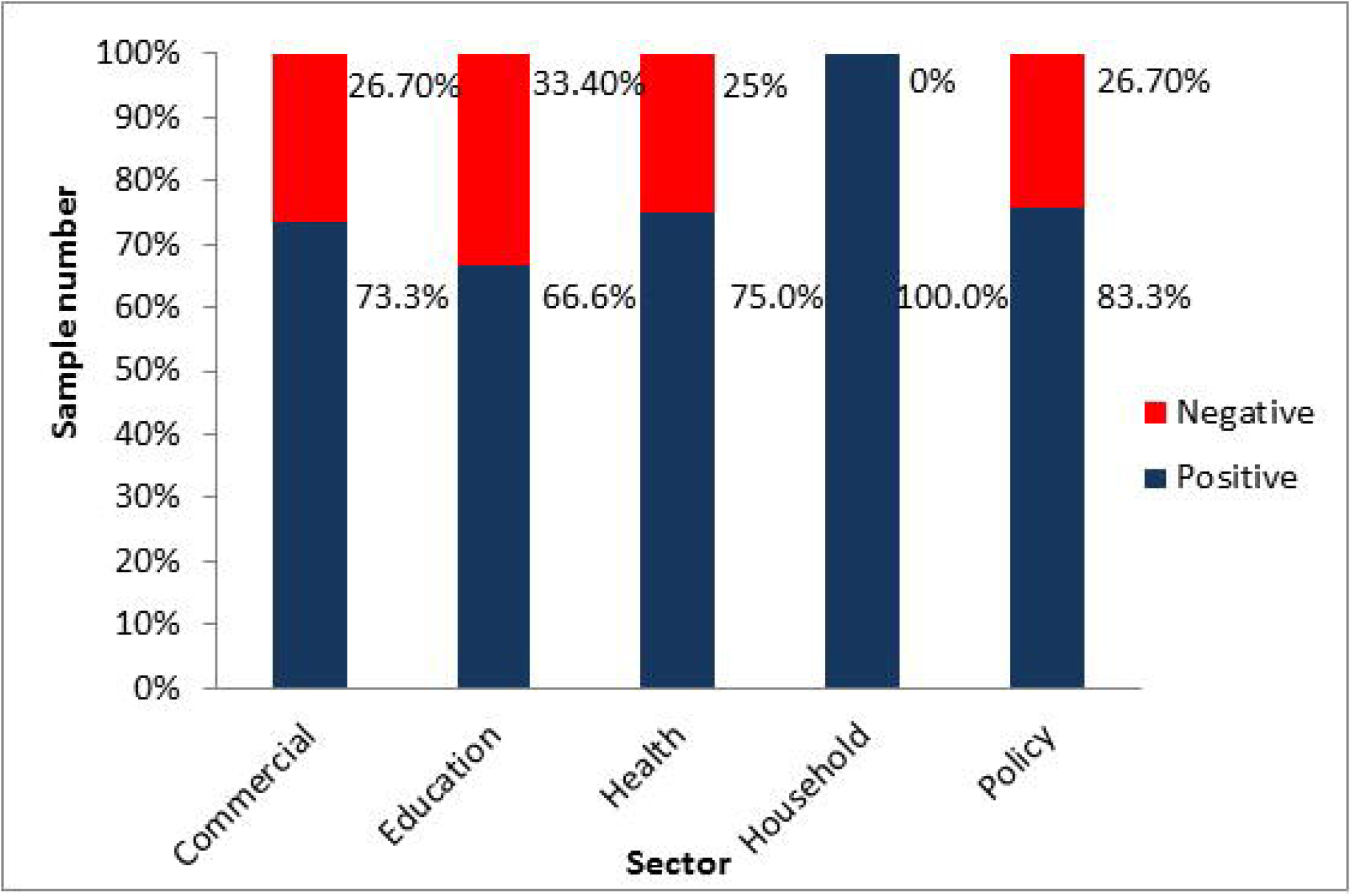

### Qualitative analysis of Quinolones residue in eggs

In a total of 120 egg samples, 96 samples (80%) were found to be positive for the residues of Quinolones group of antibiotics. Out of the 96 positive samples, 3 samples (3.12%) were found to have the residues above the MRL (i.e. 100μg/kg) (Table 3).

**Table 3:** Observed values of the positive egg samples.

| Observed values | No. of samples | Remarks |
| --- | --- | --- |
| 0 ppb | 24 | Negative |
| 1-50 ppb | 56 | Positive, since values are below the MRL value, no HPLC was performed. |
| 51-100 ppb | 37 |  |
| ≥100 ppb | 3 | Positive, since values are above the MRL value, HPLC was performed |

### Quantitative analysis of meat and eggs

The samples which were observed to have the residues above MRL value by ELISA were further quantified by HPLC method. Out of 106 positive meat samples, 9 samples (8.5%) had values equal to or above the MRL. Similarly, out of 96 positive egg samples, 3 samples (3.1%) were found to have values equal to or above the MRL. Therefore, 9 meat and 3 egg samples were analyzed through HPLC for the two mostly used antimicrobials of Quinolones group i.e. Enrofloxacin and Ciprofloxacin. Out of 9 meat samples, 5 samples were found to have residues of Enrofloxacin of which 3 samples had residues above the MRL (i.e. 100 μg/kg). Interestingly, all the 3 samples which had above the MRL were from Kathmandu district, representing one sample from commercial, education, and health sector each. Likewise, 2 samples were found to have residues of Ciprofloxacin of which one sample had residue above the MRL. The sample above the MRL belonged to the health sector from Kathmandu (Table 4). It was observed that meat samples collected from Lalitpur and Bhaktapur had no residues above the MRL value.

**Table 4:** HPLC results for positive meat sample with residues above MRL value.

| S.N. | Sector | District | Observation | Enrofloxacin (µg/kg) | Ciprofloxacin (µg/kg) |
| --- | --- | --- | --- | --- | --- |
| 1 | Commercial | Kathmandu | ≤ 1 ppm | 387.3 | ND |
| 2 | Education | Kathmandu | ≤ 1 ppm | 1120.3 | ND |
| 3 | Health | Kathmandu | ≤ 1 ppm | 246.3 | 204.2 |

ND: Not Detectable

However, out of 3 egg samples analyzed by HPLC method, all the egg samples were found to be negative for Enrofloxacin and Ciprofloxacin residues.

## DISCUSSION

The present study was conducted to assess Quinolones residues in poultry meat and eggs found in Kathmandu valley. Though Colistin sulphate was found to be the most widely used antimicrobials in both broilers and layers from the questionnaire survey, it was not chosen for the analysis. Instead, Quinolones group of antimicrobials was chosen because many poultry growers were found using the Quinolones group of antibiotics for rapid augmentation of chickens and to decrease the occurrence of diseases. The usage of these antibiotics, particularly Quinolones, has increased in the poultry industry, as supported by the survey results of this study. In addition, the withdrawal period of Colistin sulphate in meat is only 1 day and egg is zero days, while that of Enrofloxacin and Ciprofloxacin (Quinolone group) is 8-9 to 14 days. On the other hand, World Health Organization (WHO) classified the antimicrobials into three categories: critically important, highly important, and important. Quinolones group antibiotics come under critically important. Moreover, under critically important, Quinolones antibiotics belong to the highly prioritized critically important drugs (WHO, 2011). The most important ones are that Quinolones are extensively used in human medication too. And we couldn’t find any laboratories in Nepal where analysis of Colistin sulphate could be done by HPLC in meat and eggs. Therefore, we focused on analysis of Quinolones from those samples. The uncontrolled and unlimited use of these antibiotics may lead to the accumulation of undesirable residues in the animal tissues and their products which ultimately lead to emergence of anti-quinolones resistance pathogens in both poultry and clinical settings.

Quinolone residues are highly heat-stable and may persist after treatment, so the concern for residues is significant for processed and cooked meat and eggs. For example, a study reported that Enrofloxacin concentrations in chicken meat did not change after boiling or microwaving, and residues were displaced from the meat to the water (Lolo et al., 2006). However, the same study showed that roasting or grilling the meat increased the concentration of Enrofloxacin due to moisture removal during cooking.

In this study, 106 (88.33%) chicken meat samples were found positive for Quinolone antibiotic residue which is comparatively higher than that found in a similar study conducted in Ankara, Turkey, where only 45.7% of chicken meat sample were found out to be positive for Quinolones (Er et.al., 2013). The study conducted in 2007 in Kathmandu and Chitwan detected residues of Fluoroquinolones in very less proportion (5.83%) than this study which may be attributed to a lesser use of Fluoroquinolone group of antibiotics at that period of time (Pandey et. al., 2009). The higher level of residue in this study might be due to an increased as well as a haphazard use of antibiotics by farmers and poultry industry, without proper advice and prescription of the veterinarians, and poor husbandry system. Reluctance or less awareness among poultry entrepreneurs about the antibiotic withdrawal period could also be another important factor contributing to the presence of antibiotic residue in the meat samples.

While the chicken meat samples obtained from Bhaktapur had a slightly higher prevalence of Quinolone residue (92.3%) compared with that from Kathmandu (88.5%) and Lalitpur (85%), there was no statistically significant variation in occurrence of antibiotic residue in the meat samples from three districts. This suggests that the chicken meat in Kathmandu, Bhaktapur and Lalitpur contained antibiotic residues in similar proportions. Similarly, meat samples collected from all the sectors had high antibiotic residue with no significant difference, implying that all sectors were equally affected by this problem.

All the three quantified chicken meat samples belonged to Kathmandu district, one each from commercial (Enrofloxacin - 387.3 μg/kg), education (Enrofloxacin - 1120.3 μg/kg) and health sector (Enrofloxacin - 246.3 μg/kg, Ciprofloxacin - 204.2 μg/kg). The quantity of antibiotic residue detected in this study was much higher than that obtained by Sahu & Saxena (2014) in New Delhi, where Fluoroquinolones (Enrofloxacin and Ciprofloxacin) were detected in the range of 3.37 – 131.75 μg/kg. Similarly, the quantity of antibiotic residue found in chicken meat sample in the study by Er et al. (2013) was also lesser than that in our study with mean levels (±SE) of Quinolones at 30.81±0.45 μg/kg. While this difference may be due to either haphazard use of Enrofolxacin and Ciprofloxacin or slaughtering of treated birds without considering the withdrawal period, a study with a higher sample size may be needed in Kathmandu Valley for more accurate and generalizable results.

Residues in eggs may be produced by administration of antibiotics to laying hens via feed or drinking water used by veterinarians or farmers for therapy, prophylaxis and growth promotion (Herranz et al., 2007). Specifically, if Enrofloxacin is delivered by intramuscular injection to laying hens, residues can be detected in eggs after 48 hours of administration which can persist in yolk and albumen for 9 days after withdrawal (Herranz et al., 2007; Lolo et al., 2005; Maxwell et al., 1999; McReynolds et al., 2000). When Enrofloxacin is delivered in the water, higher concentrations of the residue persist in the egg than when the drug is delivered intramuscularly (Lolo et al., 2005). Furthermore, the secondary metabolite Ciprofloxacin persists for 7 to 10 days, dependent on route of administration.

In this study, 80% of the chicken egg samples were found positive for Quinolone antibiotic residue. The prevalence is higher than that found in Dominican Republic (Moscoso et al., 2015) or Sudan (Hind et al., 2014) where only 51% and 49.6% eggs collected contained Quinolone residue, respectively. This higher prevalence in this study is indicative of the haphazard use of antibiotics by Nepalese farmers, lack of knowledge about the withdrawal period, and poor husbandry practices. This is further supported by the study finding that antibiotic residues in egg samples in all three districts were of similarly high proportions. Furthermore, most of the farmer respondents believed that there was no relation between the use of antibiotics and their presence in food and eggs, and were found selling eggs even during drug administration to avoid any economic loss. In addition, lack of governmental regulations regarding egg specification may be among the other factors leading to the appearance of antibiotics in egg in this study.

In contrast, significantly more egg samples from the household sector contained Quinolone residue than those from the commercial, education, and health sector. This may be due to the difference in source from where the eggs were collected as the level of antibiotics used may be different according to source, but this can be a potentially alarming finding. However, only 3 samples were found to have residue at the MRL or above and none of them were found to be positive for Enrofloxacin and Ciprofloxacin residues.

## CONCLUSIONS

The results obtained from the questionnaire survey showed Colistin Sulphate as the most commonly used antibiotic in chicken, followed by Quinolones. A high proportion of both meat and eggs of chicken (80% or above) were found to be positive for residue of Quinolones. However, only about 2.5% of the meat samples and none of the egg samples had the residues of Enrofloxacin & Ciprofloxacin above the MRL. The antimicrobial residues on those foods not only alarms the direct impact to human health but also increases the chance of emergence of antimicrobial resistant pathogens in poultry and clinical settings. Hence, our study suggests all stakeholders to strictly stop the haphazard use of antibiotics as feed additives or for treatment. Moreover, we also recommend to aware about the withdrawal period of the antibiotics before marketing and their potential effect/s on human health.

## Data Availability

All data produced in the present study are available upon reasonable request to the corresponding and principal authors.

## Abbreviations

BMD: Bacitracin Methylene Disalicylate
CDC: Centers for Disease Control and Prevention
DNA: Deoxy-ribose Nucleic Acid
ELISA: Enzyme Linked Immuno Sorbent Assay
HPLC: High Performance Liquid Chromatography
MRL: Maximum Residue Limit
NHRC: Nepal Health Research Council
OD: Optical Density
SE: Standard Error
USP: United State Pharmacopeia
UV: Ultra Violet
WHO: World Health Organization

## Declarations

### Ethics approval and consent to participate

Written informed consent was obtained from each of the participants involved in the research study using a standard consent form. Ethical approval for this study was reviewed and granted by the Nepal Health Research Council (NHRC), Kathmandu, Nepal.

### Consent for publication

Not applicable

## Availability of data and materials

The dataset used and/or analyzed during the current study are available from the corresponding author on reasonable request.

## Competing interests

The authors declare that they have no competing interests.

## Funding

The source of funding is Feed the Future Innovation Lab of United States Agency for International Development (USAID), through Colorado State University, USA.

## Authors’ contributions

All authors had substantially contributed to the conception and design of the study, acquisition of data, or analysis and interpretation of results and equally took part in drafting the manuscript or revising it. All authors had read and approved the final version of the manuscript and agreed to submit it to this journal.

## Acknowledgements

The authors sincerely thank to Feed the Future Innovation Lab of United States Agency for International Development (USAID) and Colorado State University, USA for financial support of the work, the laboratory staff of ZEST lab for their technical support in the laboratory analyses and Prof. Dr. Richard Bowen and Dr. Diana Fahrenbruck for guiding throughout the study. Lastly, we would like to thank to Dr. Biswash Sharma for his support in data analysis.

## Notes

### Competing Interest Statement

The authors have declared no competing interest.

### Funding Statement

The study was funded by Colorado State University however, we don't have any funds to publish this manuscript.

### Author Declarations

Ethical approval for this study was reviewed and granted by the Nepal Health Research Council (NHRC), Kathmandu, Nepal

